# Drug Safety Agents Using Graphs and Ontologies

**DOI:** 10.64898/2026.02.04.26345582

**Authors:** Clint Solomon Mathialagan, Alexander Nip, Ashwani Bhat

**Affiliations:** Oracle America Inc.

## Abstract

In pharmacovigilance, analyzing drug safety cases is often time consuming due to the abundance of laboratory data, complex medical histories, and intricate temporal relationships. Agentic AI systems can significantly reduce case processing time by assisting medical reviewers in surfacing clinically relevant evidence. However, previous studies have highlighted that large language models alone lack causal reasoning and evidence-based interpretability.

To address these limitations, we present a knowledge-grounded safety case analysis framework that integrates disproportionality analysis to generate and prioritize potential adverse event hypotheses. Crucially, we introduce a novel hallucination-risk-aware execution planner that dynamically routes queries to the safest reasoning pathway, prioritizing deterministic graph retrieval over generative methods for clinically sensitive signals. The system demonstrates how structured medical knowledge and statistical evidence can be combined to support a reliable, explainable case assessment and can be readily extended with causal inference modules for deeper clinical reasoning.

## Introduction

Safety case reviewers in pharmacovigilance are often inundated with reports that lack informative content (Jarow et al. 2016), accompanied by large volumes of laboratory and historical data that are difficult to interpret efficiently. The resulting cognitive load, coupled with the high stakes of medical decision-making, contributes to significant reviewer fatigue and slower case processing.

The advent of large language models (LLMs) has created new opportunities for using AI to streamline pharmacovigilance workflows. However, several studies (Turpin et al. 2023a; Schaeffer, Miranda, and Koyejo 2023a; Kambhampati et al. 2024) have questioned the trustworthiness and reasoning reliability of such models, raising concerns about their applicability in regulated domains with high public impact.

In this work, we introduce a knowledge-grounded agentic system designed to assist in safety case analysis. Our system enhances transparency and explainability by grounding system responses in knowledge graphs and safety signal evidence, while also supporting extensions with domain-specific causal graphs to improve reliability and trust in automated pharmacovigilance.

## Related Work

Our work situates itself at the intersection of large language model (LLM) faithfulness, neuro-symbolic reasoning, and autonomous agents in healthcare.

### LLM Hallucination and Reasoning in Medicine

While LLMs have demonstrated transformative potential in biomedical tasks, their deployment in high-stakes domains is hindered by stochastic hallucinations and unfaithful reasoning. (Turpin et al. 2023b) demonstrated that chain-of-thought explanations generated by LLMs often fail to reflect the model’s actual internal decision process, creating a “plausibility illusion” that is particularly dangerous in clinical settings. Furthermore, (Schaeffer, Miranda, and Koyejo 2023b) argue that many perceived “emergent” reasoning capabilities of LLMs may be mirages of metric selection, casting doubt on their ability to perform reliable causal inference in pharmacovigilance without external grounding. Our framework addresses these fundamental limitations by decoupling *reasoning* (via the Execution Planner) from *knowledge retrieval* (via the Knowledge Graph), ensuring that safety signals are grounded in curated evidence rather than parametric memory.

### Retrieval-Augmented Generation (RAG) and Knowledge Graphs

To mitigate hallucination, Retrieval-Augmented Generation (RAG) has become the de facto standard. However, standard “naive RAG”, which retrieves unstructured text chunks, lacks the structured reasoning required for drug safety (e.g., distinguishing between a drug *treating* a symptom vs. *causing* it). Recent approaches in *GraphRAG* have sought to bridge this gap by using knowledge graphs (KGs) to enforce structural constraints on generation. In the biomedical domain, (Faruk 2025) demonstrated the utility of large-scale KGs for drug repurposing. Our work extends this paradigm by integrating *statistical signal detection* (DuMouchel and Pregibon 2001) directly into the graph schema, allowing the system to query not just for semantic relationships, but for statistically significant adverse event signals.

### Agentic AI in Healthcare

The transition from passive chatbots to active “Agents” is a rapidly evolving frontier. (Kambhampati et al. 2024) argued that while LLMs cannot truly “plan” in the algorithmic sense, they can serve as heuristic drivers for external planning modules. Recent works on medRxiv have operationalized this in clinical settings. For instance, “SAFE-AI” (Trujillo et al. 2025) proposed an ontology-driven agent to detect medication errors using strict rule-based guardrails. Similarly, (Steinbuch et al. 2026) evaluated agentic frameworks for patient history taking.

However, existing medical agents typically rely on static execution flows, either always using tools or always generating text. Our framework introduces a novel contribution to this landscape: a Hallucination-Risk-Aware Execution Planner. Unlike (Trujillo et al. 2025), which enforces rigid rules, our planner models the *probability* of hallucination as a function of query sensitivity, allowing the system to dynamically trade off between the flexibility of LLM generation and the safety of deterministic graph retrieval.

### System

Figure 1 illustrates the architecture of the agentic safety case analysis system. Each user request is processed by a query understanding agent, which interprets dialog and user interface context to extract the intent and relevant clinical entities. These entities are then resolved to corresponding nodes in the knowledge graph through an entity resolution agent that performs semantic vector search.

**Figure 1.**
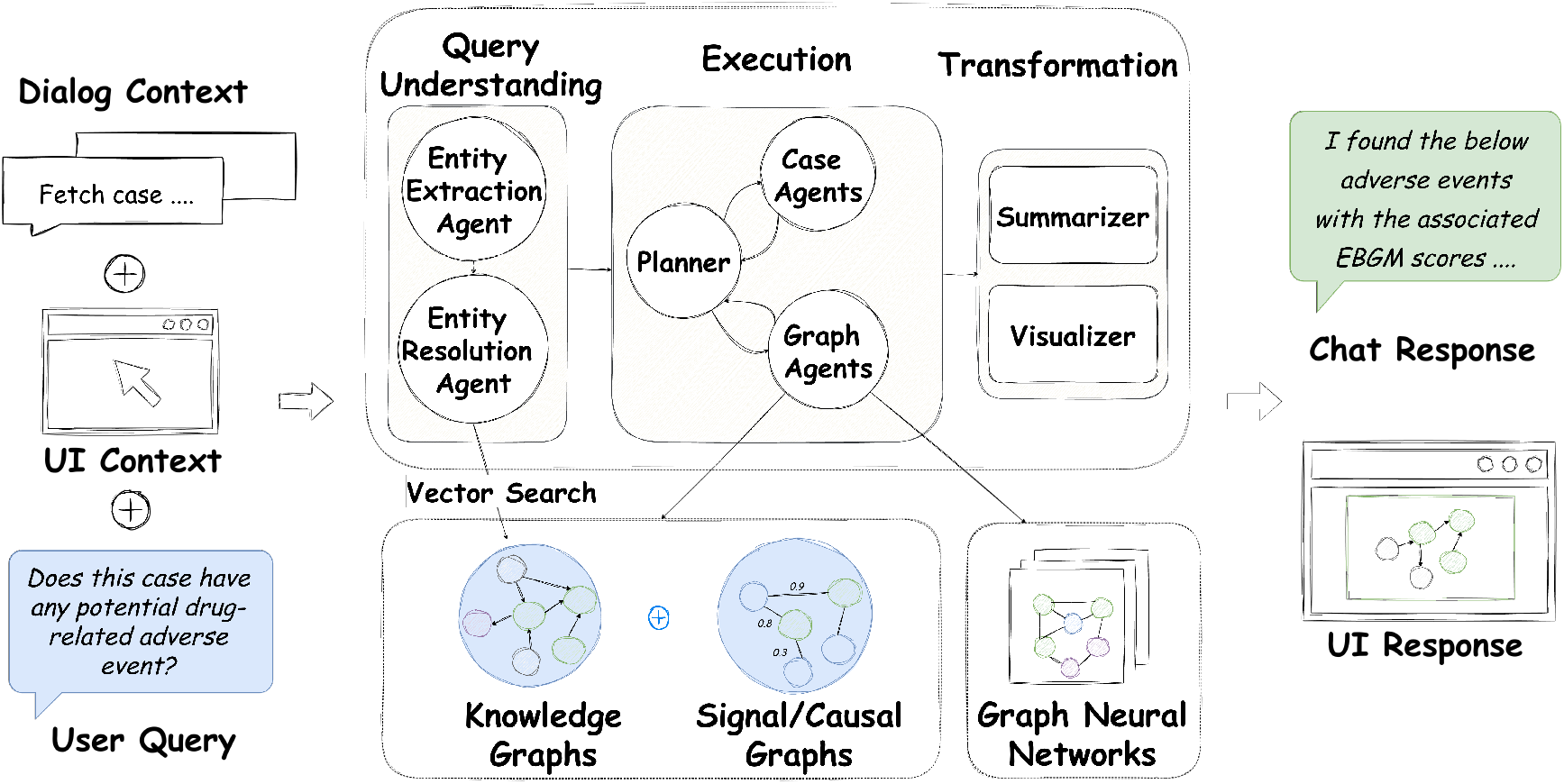
Overview of System Architecture

Using the resolved entities and identified intent, an execution plan is dynamically generated by the graph database and supporting data-retrieval agents. The retrieved information is subsequently summarized and transformed into structured graph objects by downstream transformation agents, which render the final visualizations on the user interface.

The system is implemented using Oracle Generative AI, (gen 2025) Oracle Graph Database, (gra 2025) and Oracle Graph Engine (PGX) (pgx 2025) as the core infrastructure, enabling scalable query execution and knowledge-grounded reasoning.

### Query Understanding

The query understanding subsystem functions as the entry point for risk assessment, transforming raw user input and dialog context into structured, executable intent. This proscess occurs in three stages:

#### 1. Intent and Parameter Recognition

An LLM-based extraction agent analyzes the user’s prompt to classify the primary analytical goal (e.g., Signal Detection, Causality Assessment) and extract temporal or demographic constraints. 2. **Entity Resolution and Grounding:** Extracted clinical mentions (drugs, symptoms) are resolved to canonical nodes in the Knowledge Graph (aligned to MedDRA) using semantic vector search. To support multi-turn conversations, the agent resolves coreferences (e.g., mapping “that drug” to the entity focused on in the previous turn). 3. **Ambiguity Metadata Generation:** Crucially, the resolution agent calculates quantitative metadata. These metrics are passed downstream to the Query Ambiguity Classifier to help determine the hallucination risk profile of the request before execution begins.

### Execution

After entities and parameters are resolved, the execution planner uses LLM agents to determine the appropriate reasoning pathway (Figure 2). The planner may employ one of the below strategies:

**Figure 2.**
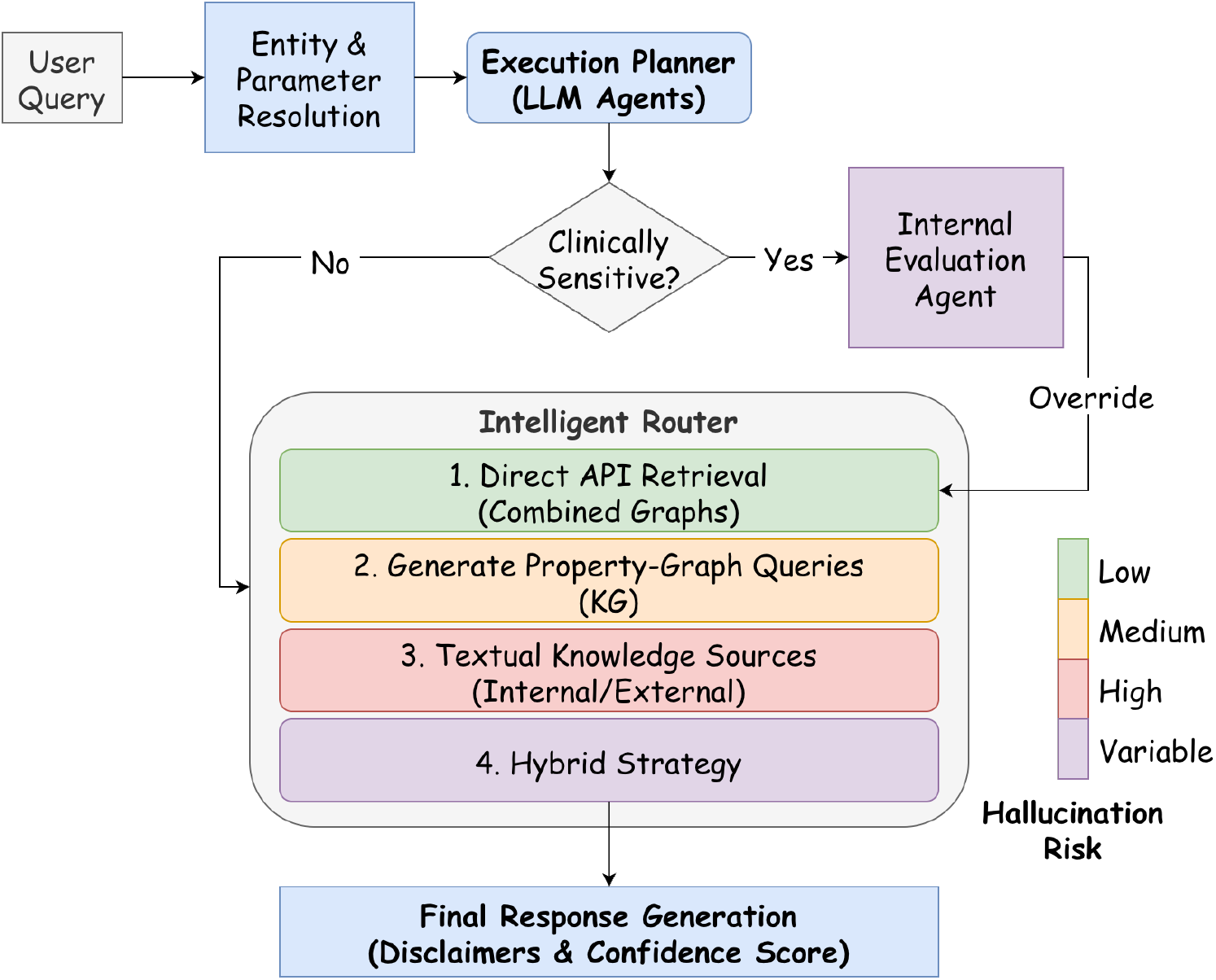
Hallucination Mitigation Framework

**Figure 3.**
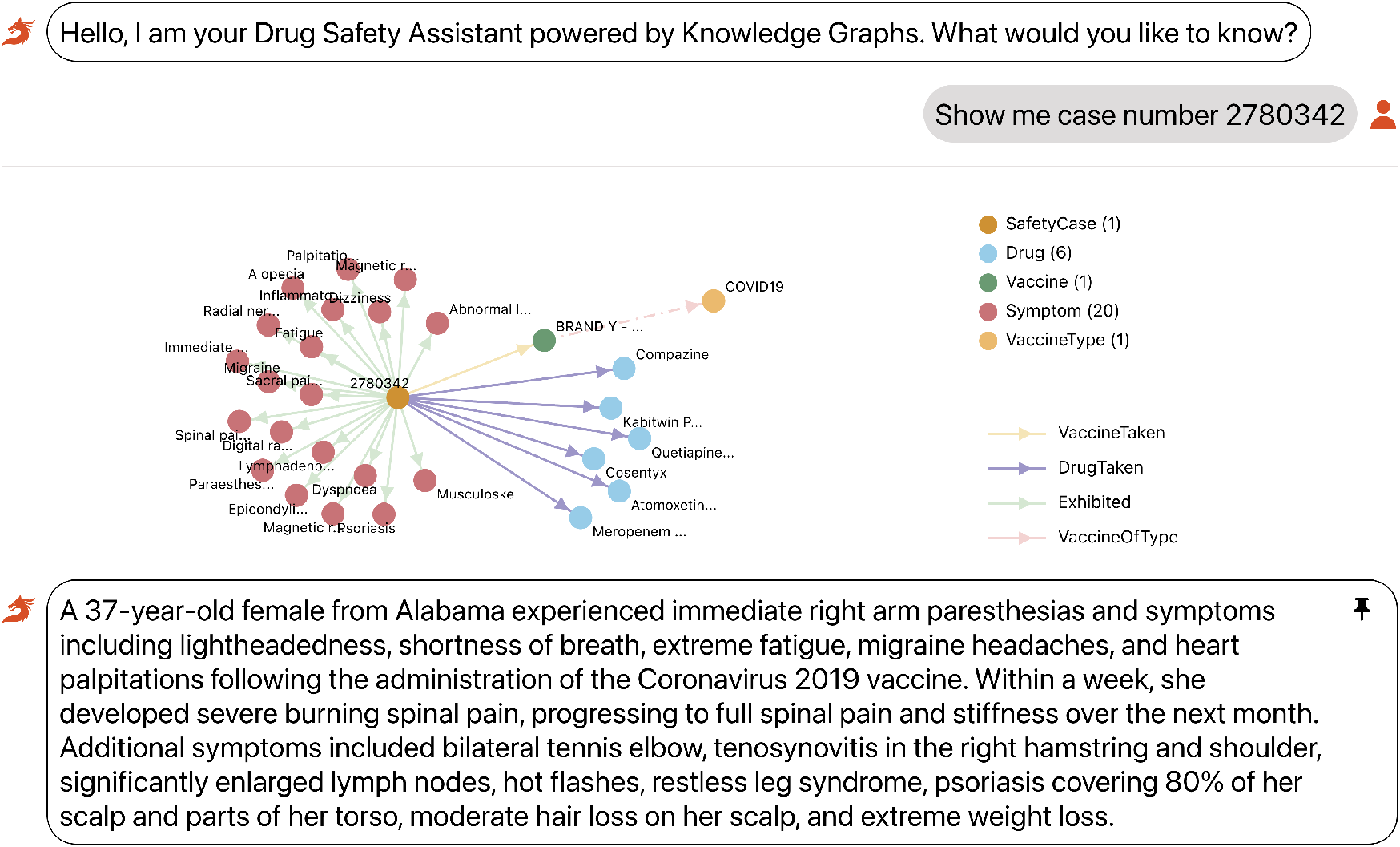
User Interacting with the System

1. call an API that retrieves results directly from the combined knowledge and signal graphs;
2. generate property-graph queries that operate over the KG;
3. respond using external or internal textual knowledge sources; or
4. employ a hybrid strategy involving the above and specialized models like a GNN.

We categorize these execution strategies by hallucination risk: direct evidence retrieval (1) is lowest risk, KG-based query generation (2) carries moderate risk, and free-form knowledge-based responses (3) carry the highest risk. For clinically sensitive queries, such as those involving causality or elevated risk signals, the system prioritizes low-risk strategies and only escalates to higher-risk pathways when the internal evaluation agent determines that lower-risk routes cannot satisfy the request. The final resolved pathway is recorded and used to generate appropriate disclaimers and assign a conservative confidence score.

### Transformation

The transformation layer serves as the final interpretability bridge, converting complex graph query results into verifiable clinical insights. This process executes in two parallel streams to ensure that every generated claim can be visually audited by the user.

### Visualizer

Raw outputs from the execution layer (e.g., time-series data from signal graphs or subgraph isomorphism results) are first serialized into a standardized intermediate JSON representation. These objects are rendered as interactive widgets, such as Gantt charts for event timelines or force-directed graphs for drug interactions (Figures 4, 5).

**Figure 4.**
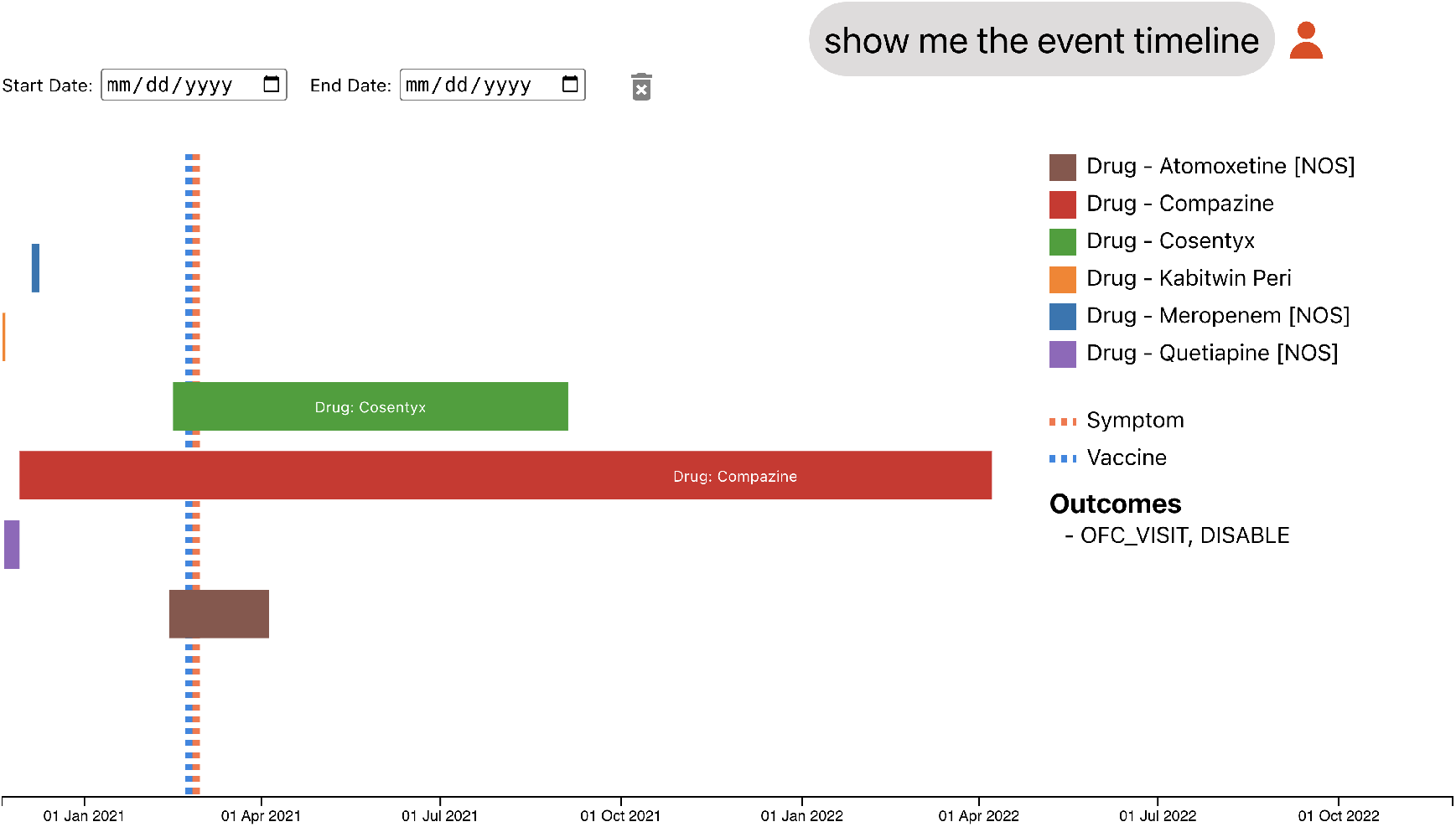
User Inspecting Case Timeline

**Figure 5.**
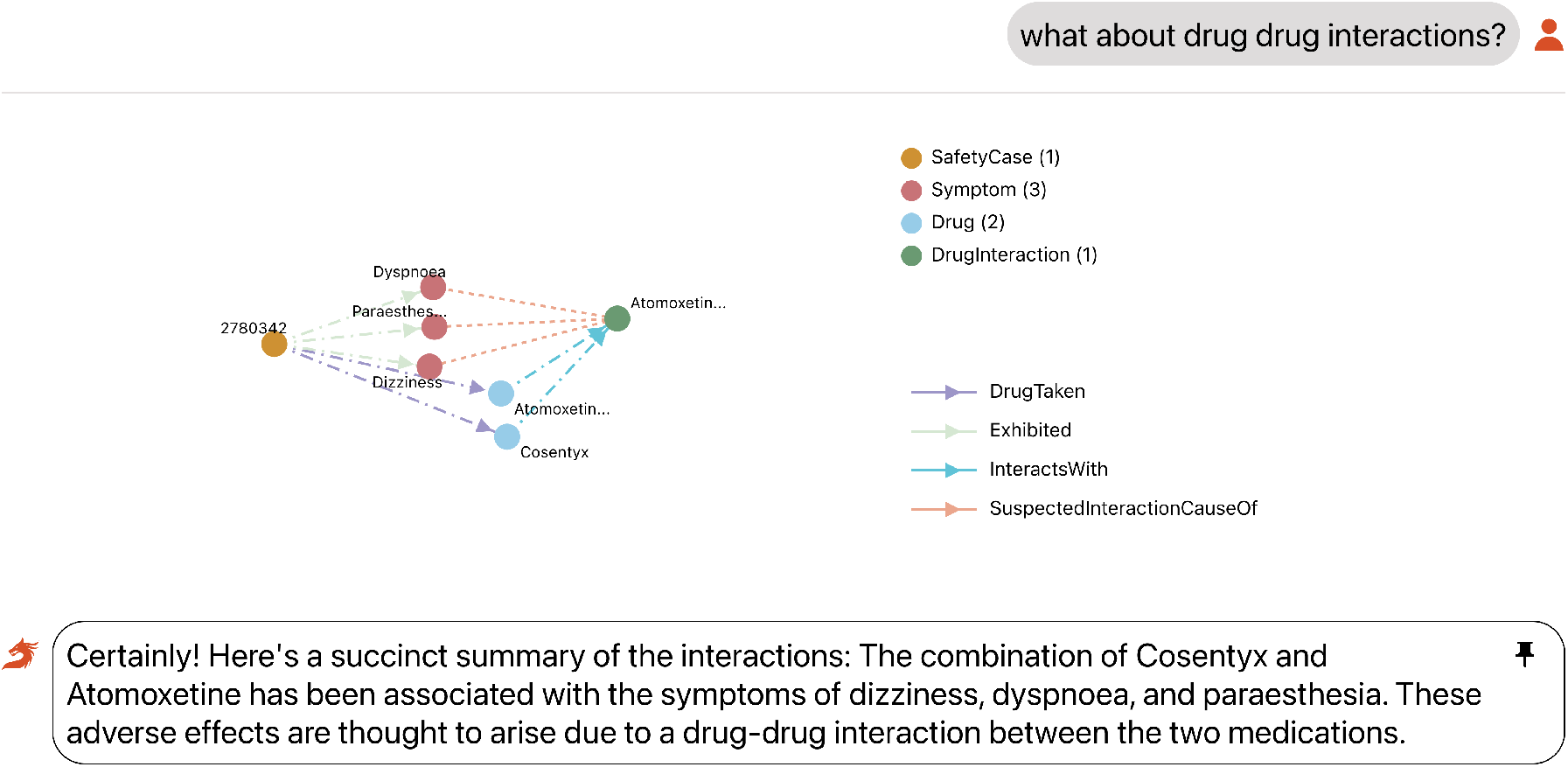
System Responding with Evidence

Critically, these visualizations are not static images; every node and edge is hyperlinked back to its source evidence. For example, clicking a “Drug-Event” edge allows the medical reviewer to drill down into the specific statistical signal scores (EBGM/INTEB) that support the association, ensuring complete data lineage.

### Summarizer

Simultaneously, an LLM summarizer generates a natural-language narrative of the findings. Unlike the upstream reasoning agents, this model operates under strict *extractive constraints*, forcing it to cite specific nodes from the retrieved subgraph rather than generating free text.

Presenting visual data alongside cited text enables a “human-in-the-loop” verification workflow. Users can independently validate the narrative summary against the interactive visual evidence, significantly mitigating the risk of undetected hallucinations in the final report.

## Hallucination-Risk-Aware Execution Planning

### Definitions and Space

We model the Execution Planner (as shown in (Figure 2)) as a constrained optimization problem over a discrete action space. Let *Q* be the space of all possible user queries. Let 𝒮 be the set of *k* available execution strategies, ordered by their intrinsic risk profile:

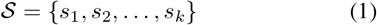

where *s*_1_ represents the most deterministic strategy (e.g., direct API invocation) and *s*_*k*_ represents the most generative strategy (e.g., free-form LLM generation).

### Oracles and Functions

We define three key functions acting on a query *q* ∈ *Q*:

1. Sensitivity Function (Clinical Guardrail)

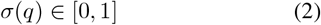

Where *σ*(*q*) → 1 denotes maximum clinical sensitivity (e.g., causality, adverse events) and *σ*(*q*) → 0 denotes benign information seeking.
2. Hallucination Oracle

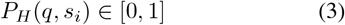

This represents the estimated probability that strategy *s*_*i*_ will produce a hallucination for query *q*. We assume a monotonic increase in risk across the ordered set 𝒮:

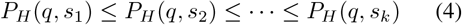
3. Evaluation Agent (Utility Function)

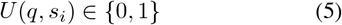

A binary outcome from the internal evaluation agent, where 1 indicates the strategy *s*_*i*_ successfully retrieved a valid answer and 0 indicates failure or empty retrieval.

### Optimal Strategy

The planner’s objective is to select an optimal strategy *s*^∗^ that satisfies the user’s information need while minimizing risk, constrained by the query’s sensitivity.

We define a **Risk Threshold Function** *τ* (*σ*), which is inversely proportional to the query sensitivity. As clinical sensitivity *σ*(*q*) increases, the acceptable risk tolerance decreases:

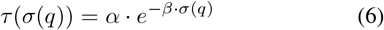

where *α* and *β* are system-specific tuning constants.

The planner searches for the optimal strategy *s*^∗^ by iterating through the set 𝒮 to find the lowest-indexed strategy that satisfies both utility and safety conditions:

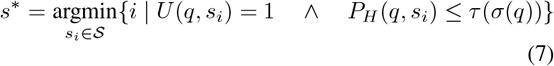

The final system confidence *C*_*sys*_ for a response generated by strategy *s*_*i*_ is modeled as:

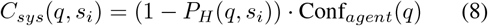

If no strategy satisfies the safety condition (i.e., ∀*i* ∈ {1, …, *k}, P*_*H*_ (*q, s*_*i*_) *> τ* (*σ*(*q*))), the system enters a failure mode, returning a safe fallback response *R*_*fallback*_.

### Implementation

To instantiate the theoretical framework defined above, we approximate the idealized oracles using lightweight, commercially viable classifiers.

#### Approximating the Sensitivity Function *σ*(*q*)

We implement the sensitivity function *σ*(*q*) using a transfer learning approach tailored to biomedical text. Instead of training a model from scratch, we utilize PubMedBERT (Gu et al. 2020) to generate dense vector embeddings for the input query. These embeddings are fed into a lightweight MultiLayer Perceptron (MLP) classification head.

The classifier was trained on a labeled dataset of pharmacovigilance queries to predict a scalar probability 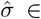 [0, 1]. By freezing the pretrained PubMedBERT backbone and only training the classification head, we leverage the model’s deep semantic understanding of medical terminology (e.g., distinguishing “adverse event” contexts from general mechanism-of-action queries) while maintaining computational efficiency. The resulting score 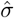 serves as the direct input to our dynamic threshold function *τ* .

#### Approximating the Hallucination Risk *P*_*H*_

Since a perfect hallucination oracle is unattainable, we estimate *P*_*H*_ (*q, s*_*i*_) using a *proxy risk model*. We assign a static base risk score *R*_*base*_(*s*_*i*_) to each strategy derived from historical error rates on a hold-out validation set (e.g., *R*_*base*_(*s*_1_) = 0.02, *R*_*base*_(*s*_*k*_) = 0.45). This base risk is dynamically modulated by a “Query Ambiguity Classifier”, a Random Forest model. Critically, this classifier is not trained on raw text, but on *entity-resolution metadata*.

Features include:

##### 1. Entity Grounding Score

The aggregate confidence score from the entity linking step (e.g., linking “Advil pills” to corresponding Drug node in the Knowledge Graph).

##### 2. Ontological Specificity

The average depth of resolved concepts in the Knowledge Graph hierarchy (detecting vague terms like “drugs” vs. specific terms like “acetaminophen”).

##### 3. Unresolved Token Ratio

The percentage of substantive tokens that failed to map to any known medical entity.

This design ensures that “ambiguity” is defined not by linguistic complexity, but by the lack of grounded signal in the Knowledge Graph.

##### Execution Logic

The optimization loop is implemented as a deterministic “waterfall” control flow. The system iterates through strategies *s*_1_ … *s*_*k*_. For each *s*_*i*_, the Utility function *U* (*q, s*_*i*_) is realized as a null-check on the retrieval result (i.e., did the API return non-empty data?). If *U* = 1 and the estimated risk is below the calculated threshold 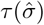, the loop terminates, and the result is served. If the loop exhausts all strategies without satisfying the safety constraint, the fallback response is triggered.

## Data and Evaluation

### Synthetic Data

To prototype the system, we constructed a knowledge graph from a synthetic dataset derived from curated subsets of the publicly available VAERS and FAERS databases (fae 2025), aligned to MedDRA terminology (med 2025). Disproportionality-analysis metrics, including the empirical Bayes geometric mean (EBGM) and the posterior limit of the interaction estimate (INTEB) (DuMouchel and Pregibon 2001), were computed to construct signal graphs and quantify association strength.

Users interact with the system through a chat-based interface (Figures 3, 4, 5). By issuing natural-language queries such as “Show me cases with elevated liver enzymes after drug X” or “Find vaccines associated with increased myocarditis reports in young adults,” users can explore temporal patterns, entity relationships, and evidence links within the knowledge graph. The interface highlights relevant entities, surfaced hypotheses, and disproportionality metrics, enabling rapid contextualization of safety findings.

### Evaluation

We conducted a preliminary hallucination-focused evaluation of the lowest-risk execution mode (strategy 1). We curated an evaluation set consisting of:

1. 50 drug–symptom pairs with strong, known evidence;
2. 50 drug–random-symptom pairs where the evidence score was artificially strong; and
3. 50 drug–symptom pairs with weak evidence for known side effects.

For each of the 150 queries, we audited whether the model summary accurately reflected the retrieved evidence. In all cases, the agent faithfully grounded its responses in the underlying evidence and did not introduce unsupported hallucinations. In future work, we plan to extend evaluation to higher-risk execution pathways (strategies 2 and 3) and to assess robustness across more diverse query types.

## Limitations

While our framework mitigates large language model (LLM) hallucinations through knowledge and evidence grounding, residual risks of hallucination remain. Biases introduced by the underlying LLM or its sampling mechanisms may still surface, requiring users to exercise appropriate due diligence when interpreting generated insights.

For components employing graph neural networks, standard machine learning risks such as overfitting, representation bias, and model drift continue to apply. Additionally, the demo integrates Oracle Empirica Signal Detection as part of its analytical workflow; however, the generated associations should not be interpreted as verified causal relationships.

Finally, the demo operates on synthetic data containing intentionally exaggerated adverse events for the sole purpose of illustrating system capabilities, and does not reflect realworld pharmacovigilance findings.

## Conclusion

We believe that our work provides a robust framework for integrating AI-driven systems into pharmacovigilance workflows. In future work, we plan to expand the platform to incorporate additional data sources such as electronic health records (EHRs), clinical trial data, and biomedical literature (e.g., PubMed), as well as broader domain ontologies including SNOMED CT, LOINC, and related vocabularies.

These extensions will enable the development of comprehensive capabilities that embed safety considerations across multiple stages of the medical product life cycle—from early signal detection to post-marketing surveillance.

## Data Availability

The raw pharmacovigilance data used to generate the synthetic datasets for this study are publicly available from the U.S. Food and Drug Administration (FDA) via the FAERS and VAERS databases. The specific knowledge graph constructed for this study was derived from a synthetic subset of these databases to simulate exaggerated adverse event signals. Access to the specific synthetic dataset for the proposed framework is restricted and proprietary.

## Acknowledgments

We thank the various teams and collaborators at Oracle whose support and infrastructure made this work possible. We also acknowledge the U.S. Food and Drug Administration (FDA) for maintaining and providing access to the public pharmacovigilance data sources that enabled this research.

## Disclaimer

This work may contain forward-looking statements; the agents and workflows described are Oracle research prototypes, are not generally available products or features, were used solely for research purposes, and no commitment is made regarding future availability, functionality, or timing.

## Notes

### Competing Interest Statement

All authors are current employees of Oracle Corporation. The framework described in this manuscript utilize commercial software technologies (Oracle Graph Database, Oracle Generative AI Service, and Oracle Empirica) developed and marketed by Oracle. The authors may hold equity or stock options in Oracle Corporation. This work was conducted as part of the authors' regular employment duties.

### Funding Statement

This study did not receive any external funding

### Author Declarations

Public Data: FAERS (https://www.fda.gov/drugs/fdas-adverse-event-reporting-system-faers/fda-adverse-event-reporting-system-faers-public-dashboard) VAERS (https://vaers.hhs.gov/) Subsets of the above were use to create synthetic adverse event datasets for evaluation of the system and classifier training datasets.

## References

1. 2025. Empirica. https://www.oracle.com/life-sciences/safety-solutions/empirica-safety-signal-management/. Accessed: 2025-10-19.

2. 2025. FDA Adverse Event Reporting System (FAERS). https://www.fda.gov/drugs/fdas-adverse-event-reporting-system-faers/fda-adverse-event-reporting-system-faers-latest-quarterly-data-files. Accessed: 2025-10-19.

3. 2025. Generative AI Service. https://www.oracle.com/artificial-intelligence/generative-ai/generative-ai-service/. Accessed: 2025-10-19.

4. 2025. GraphDB. https://www.oracle.com/database/integrated-graph-database/. Accessed: 2025-10-19.

5. 2025. Medical Dictionary for Regulatory Activities (MedDRA). https://www.meddra.org/. Accessed: 2025-10-19.

6. 2025. Parallel Graph AnalytiX (PGX). https://www.oracle.com/middleware/technologies/parallel-graph-analytix.html. Accessed: 2025-10-19.

7. DuMouchel, W.; and Pregibon, D. 2001. Empirical bayes screening for multi-item associations. In Proceedings of the Seventh ACM SIGKDD International Conference on Knowledge Discovery and Data Mining, KDD ‘01, 67–76. New York, NY, USA: Association for Computing Machinery. ISBN 158113391X.

8. Faruk, M. O. 2025. A Large-Scale Pharmacogenomic Knowledge Graph for Drug-Gene-Variant-Disease Discovery. medRxiv.

9. Gu, Y.; Tinn, R.; Cheng, H.; Lucas, M.; Usuyama, N.; Liu, X.; Naumann, T.; Gao, J.; and Poon, H. 2020. Domain-Specific Language Model Pretraining for Biomedical Natural Language Processing. CoRR, abs/2007.15779.

10. Jarow, J. P.; Casak, S.; Chuk, M.; Ehrlich, L. A.; and Khozin, S. 2016. The Majority of Expedited Investigational New Drug Safety Reports Are Uninformative. Clinical Cancer Research, 22(9): 2111–2113.

11. Kambhampati, S.; Valmeekam, K.; Guan, L.; Verma, M.; Stechly, K.; Bhambri, S.; Saldyt, L. P.; and B Murthy, A. 2024. Position: LLMs Can’t Plan, But Can Help Planning in LLM-Modulo Frameworks. In Salakhutdinov, R.; Kolter, Z.; Heller, K.; Weller, A.; Oliver, N.; Scarlett, J.; and Berkenkamp, F., eds., Proceedings of the 41st International Conference on Machine Learning, volume 235 of Proceedings of Machine Learning Research, 22895–22907. PMLR.

12. Schaeffer, R.; Miranda, B.; and Koyejo, S. 2023a. Are emergent abilities of large language models a mirage? In Proceedings of the 37th International Conference on Neural Information Processing Systems, NIPS ‘23. Red Hook, NY, USA: Curran Associates Inc.

13. Schaeffer, R.; Miranda, B.; and Koyejo, S. 2023b. Are Emergent Abilities of Large Language Models a Mirage? 2304.15004.

14. Steinbuch, S.; de Vos-Hillebrand, L.; O’Neill-Dee, C.; Wu, I.; Steinbuch, H.; Kulcsár, Z.; Ranjan, A.; Verma, A.; Landsberg, J.; Dietrich, D.; Hardin, C. C.; Jain, R. K.; and Subudhi, S. 2026. Comparative Performance of agentic AI and Physicians in Taking Clinical History across Leading Large Language Models (LLMs). medRxiv.

15. Trujillo, D.; Wang, D.; Bahr, N.; Yi-Jin Hsieh, T.; Cho, B.; Meckler, G.; Hansen, M.; Eriksson, C.; Kim, K. S.; Bedrick, S.; Jiang, X.; and Guise, J.-M. 2025. A medically grounded LLM agent–based tool to detect patient safety events in medical records. medRxiv.

16. Turpin, M.; Michael, J.; Perez, E.; and Bowman, S. R. 2023a. Language models don’t always say what they think: unfaithful explanations in chain-of-thought prompting. In Proceedings of the 37th International Conference on Neural Information Processing Systems, NIPS ‘23. Red Hook, NY, USA: Curran Associates Inc.

17. Turpin, M.; Michael, J.; Perez, E.; and Bowman, S. R. 2023b. Language Models Don’t Always Say What They Think: Unfaithful Explanations in Chain-of-Thought Prompting. 2305.04388.

